# Prognostic value of artificial intelligence-derived echocardiographic measurements in transthyretin cardiomyopathy

**DOI:** 10.64898/2026.04.01.26349281

**Authors:** Amely Walser, Andreas J. Flammer, Moritz J. Hundertmark, Isaac Shiri, Nicola Ciocca, Christoph Ryffel, Stefano de Marchi, Rahel Schwotzer, Frank Ruschitzka, Felix C. Tanner, Christoph Gräni, Dominik C. Benz

**Author notes:** Email addresses. **Corresponding author:** Dominik C. Benz, MD, Department of Nuclear Medicine and Cardiology, University Hospital of Zurich, Ramistrasse 100, 8091 Zurich, CH-Switzerland.

## Abstract

**Background:** Transthyretin cardiomyopathy (ATTR-CM) is a progressive, potentially fatal disease requiring accurate risk stratification. Echocardiography is the first-line imaging modality, with AI-based tools increasingly applied for automated analysis, yet their prognostic value remains unknown.

**Objectives:** To examine the prognostic value of AI-derived echocardiographic measurements and their incremental value beyond biomarker staging in ATTR-CM.

**Methods:** This retrospective study included patients from two ATTR-CM registries. Baseline echocardiograms were analyzed using the fully automated AI-based software Us2.ai. Prognostic performance was assessed by Kaplan-Meier analysis, Cox regression, and ROC curves. A two-parameter echocardiographic staging system combining left ventricular (LV) global longitudinal strain (GLS) and right ventricular (RV) fractional area change (FAC) stratified patients into low (both normal), intermediate (one abnormal), and high risk (both abnormal).

**Results:** Among 347 patients (91% male, median age 78 years), 141 experienced all-cause death or heart failure hospitalization over a median follow-up of 2.4 years. In multivariable analysis, AI-derived LV-GLS (HR 1.13 [1.03-1.25], p=0.011) and RV FAC (HR 0.96 [0.93-0.99], p=0.014) were independent outcome predictors. Echo staging stratified risk into groups with 3-fold (95% CI 1.70-5.91) and 6-fold (95% CI 3.22-10.30) increased hazard compared to low risk (p<0.001), with incremental prognostic value beyond National Amyloidosis Centre (NAC) staging and age (chi-square from 53 to 80; p<0.001). AI and human measurements showed comparable 1-year predictive performance (all p>0.05).

**Conclusion:** AI-derived echocardiographic measurements demonstrate independent and incremental prognostic value beyond biomarker-based NAC staging in ATTR-CM, comparable to human measurements, supporting their integration into clinical risk stratification.

## INTRODUCTION

Cardiac amyloidosis (CA) is caused by the deposition of misfolded proteins as amyloid fibrils in the myocardium. Transthyretin amyloid cardiomyopathy (ATTR-CM) is the most prevalent form and occurs either as wild-type (ATTRwt) or hereditary variant (ATTRv). As untreated cardiac amyloidosis is associated with high mortality, early and accurate risk stratification is essential.

Currently, staging for ATTR is mostly based on the National Amyloidosis Centre (NAC) staging^1^ system relying on laboratory values. Growing evidence suggests that echocardiographic parameters, particularly GLS, offer prognostic value beyond NAC biomarker staging in ATTR-CM, though the evidence base continues to expand. In a very recent study including 816 patients with ATTR-CM, a novel imaging-based prognostic staging system has been proposed, based on baseline GLS assessment at the time of diagnosis of wild-type transthyretin amyloid cardiomyopathy.^2^ GLS was found to predict mortality independent of both tafamidis initiation and Gilmore National Amyloid Centre (NAC) stage, and GLS provided incremental discriminatory value within each NAC stage. With the increasing availability of AI-based automated echocardiography platforms in clinical practice, the crucial question arises as to whether AI-derived measurements retain this prognostic value. It is noteworthy that despite the rapid growth of AI in medicine, less than 2% of approved AI-enabled medical devices currently perform a prognostic function^3^, highlighting a critical gap between AI development and validated prognostic application in clinical practice. Demonstrating the prognostic value of AI-derived echocardiographic measurements would therefore not only complement existing staging frameworks but also provide important independent prognostic validation of the AI measurements themselves.

The aim of this study is to examine the prognostic value of AI-derived echocardiographic measurements obtained at diagnosis for predicting outcomes in patients with cardiac amyloidosis and to investigate whether AI-based measurements offer added value over existing biomarker-based staging systems. We hypothesize that AI-derived echocardiographic measurements have significant, independent prognostic value that could complement existing biomarker-based staging systems. Additionally, we aim to compare the prognostic performance of AI-derived versus human-measured echocardiographic parameters.

## METHODS

### Study population

This retrospective cohort study, approved by the Zurich Cantonal Ethics Committee (KEK-ZH 2014-0490) as well as the Bern Cantonal Ethics Committee (KEK: 2021-00135), included 347 patients from two registries with transthyretin amyloid cardiomyopathy (ATTR-CM). All participants provided written informed consent. Patients underwent echocardiograms between 2005-2024, performed at the University Hospitals Zurich and Bern, and less frequently, in other Swiss hospitals or private practices. For each patient, a baseline echocardiogram obtained at or shortly after diagnosis was selected for prognostic analysis. Serum biomarkers and functional status were selected within three months of this echocardiography.

### Image acquisition and analysis

All echocardiograms were analyzed using the AI-based software US2.ai, which provided fully automated quantification. Routine echocardiographic reports, performed by board-certified cardiologists, were used for comparison with AI-derived measurements.

### Assessment of Outcomes

Patient follow-up was obtained by medical charts from treating physicians (i.e., general practitioner or cardiologist) or from the hospital’s clinical information system. The primary endpoint was a composite of all-cause mortality or heart failure hospitalization, whichever occurred first. Follow-up was censored at the time of death, heart failure hospitalization, or last clinical visit.

### Disease Staging

Disease severity was assessed using the established National Amyloidosis Centre (NAC) staging system, classifying patients into Stage I (NT-proBNP ≤3,000 pg/mL and eGFR ≥45 mL/min/1.73 m²), Stage III (NT-proBNP >3,000 pg/mL and eGFR <45 mL/min/1.73 m²), and Stage II (all remaining patients with one abnormal biomarker).^1^

### Statistical Analysis

Data are reported as medians and interquartile ranges (IQR), and hazard ratios (HR) with 95% confidence intervals (CI), as appropriate. First, associations between baseline characteristics and outcomes were evaluated using uni- and multivariable Cox proportional hazards regression. To avoid immortal time bias, tafamidis treatment was evaluated as a time-dependent covariate. For the multivariable Cox regression analysis, echocardiographic variables were selected based on availability in the majority of patients, absence of strong collinearity (assessed via a correlation matrix), and statistical significance in univariable analysis. One parameter was selected per major echocardiographic domain, representing LV structure (IVSd), LV systolic function (LV-GLS), LV diastolic function (E/e’), and RV function (RV FAC). The model was additionally adjusted for age and sex. Second, event-free survival was assessed using Kaplan-Meier analysis with log-rank tests. Parameters were initially stratified using established clinical cutoffs to classify values as normal or abnormal. For abnormal values, optimal cutoffs to further stratify into mild and severe categories were determined by log-rank maximization. Third, an echo staging system was developed by systematically screening candidate echocardiographic parameters in combination with LV-GLS. LV-GLS was selected as the fixed first parameter based on its strong independent prognostic value in both univariable and multivariable analyses and its established role in cardiac amyloidosis risk stratification. Each parameter combination was evaluated using two criteria. First, incremental prognostic value beyond established biomarker-based NAC staging and age was assessed by comparing sequential Cox regression models using likelihood ratio chi-square tests (Δχ²) and C-index improvements (ΔC-index). Second, the ability to further risk-stratify patients within each NAC stage was evaluated by comparing annualized event rates between echo risk categories using Poisson regression. Parameters were ranked by a composite score: (Δχ² × 2) + (mean χ² across stages × 3) + (ΔC-index × 100). The optimal two-parameter combination (LV-GLS and RV FAC) stratified patients into low (both normal), moderate (one abnormal), and high risk (both abnormal) (**Supplemental Table S1**). Lastly, receiver operating characteristic (ROC) curves assessed 1-year event prediction, comparing AI-derived versus human-measured echocardiographic parameters. Areas under the curve (AUC) were compared using DeLong’s test. For this comparative analysis, the first echocardiogram at or after diagnosis with both measurements available was selected for analysis. All analyses were performed using R version 4.5.2 with packages tidyverse, survival, survminer, and pROC. Two-sided p-values <0.05 were considered statistically significant.

## RESULTS

### Study Population

Median age at the time of baseline echocardiography was 78 (Interquartile range (IQR), 72–82) years and 317 (91%) were male. 320 patients (92%) had wild-type, the remaining 27 had hereditary amyloidosis. Atrial fibrillation was present in 47% of patients. Echocardiography demonstrated a median interventricular septal thickness of 14 mm (IQR 12–16), a median LVEF of 55% (IQR 46–61), and a median LV-GLS of −14.4% (IQR −17.1 to −12.0). Median NT-proBNP at the time of imaging was 1,745 pg/mL (IQR 878–3,185). Baseline demographic, clinical, and laboratory characteristics are summarized in **Table 1**. Baseline echocardiographic characteristics are summarized in **Table 2**. Over a median follow-up of 2.4 years (IQR, 1.1–4.1), 141 of 347 patients (41%) experienced the composite endpoint of all-cause death or heart failure hospitalization, including 60 deaths (17%) and 81 first heart failure hospitalizations (23%).

**Table 1.**
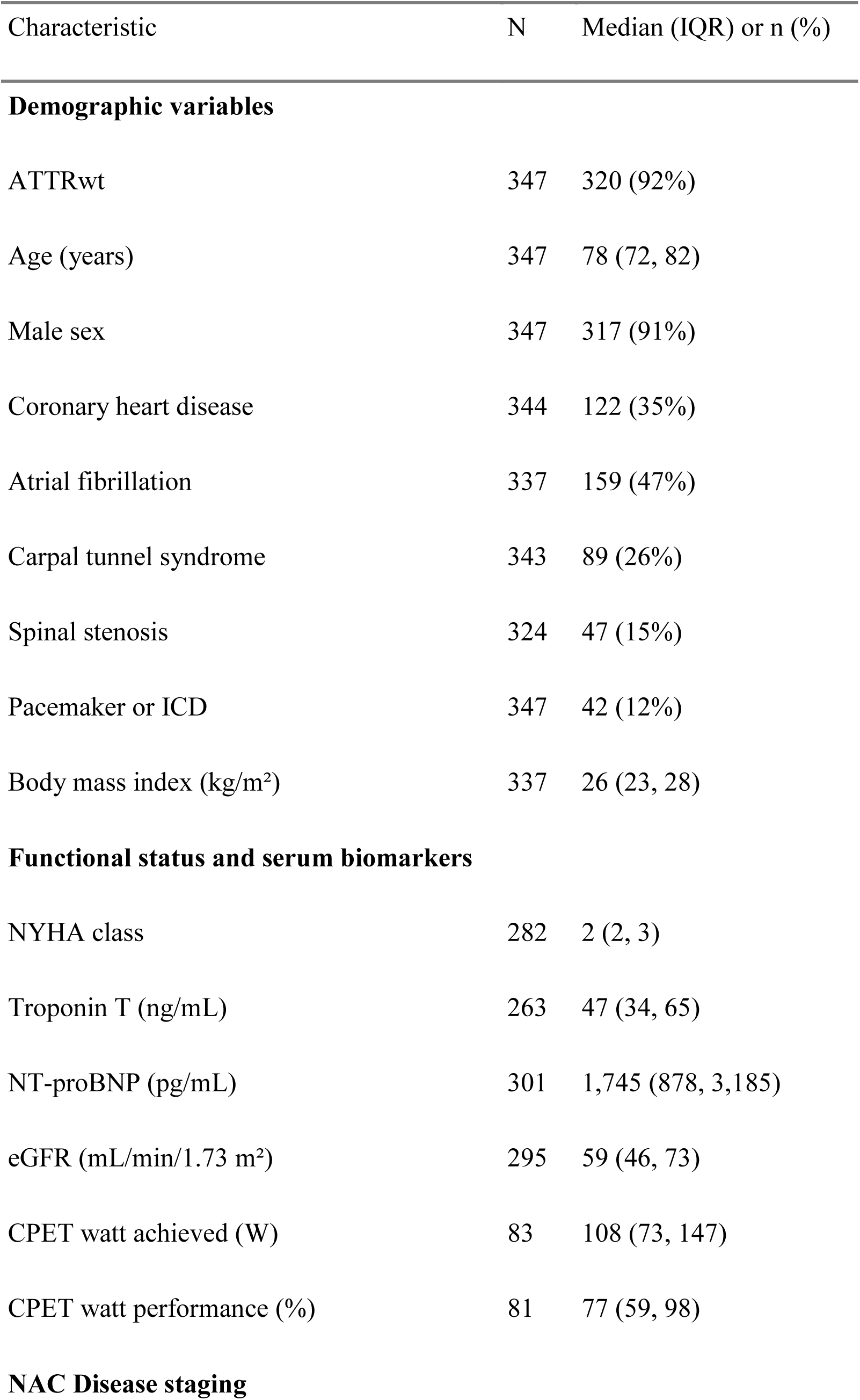
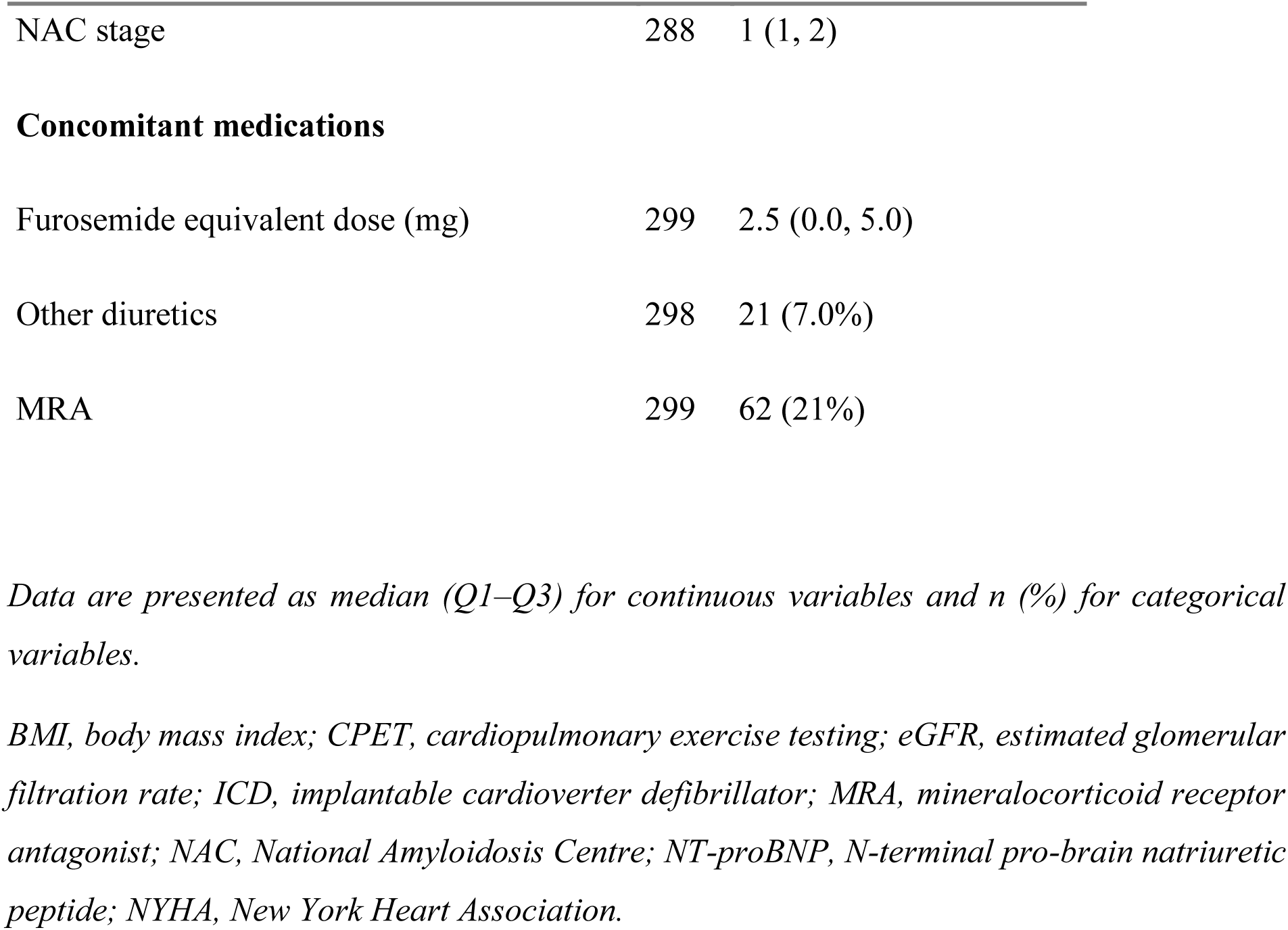
Baseline characteristics.

| Characteristic | N | Median (IQR) or n (%) |
| --- | --- | --- |
| **Demographic variables** |  |  |
| ATTRwt | 347 | 320 (92%) |
| Age (years) | 347 | 78 (72, 82) |
| Male sex | 347 | 317 (91%) |
| Coronary heart disease | 344 | 122 (35%) |
| Atrial fibrillation | 337 | 159 (47%) |
| Carpal tunnel syndrome | 343 | 89 (26%) |
| Spinal stenosis | 324 | 47 (15%) |
| Pacemaker or ICD | 347 | 42 (12%) |
| Body mass index (kg/m<sup>2</sup>) | 337 | 26 (23, 28) |
| **Functional status and serum biomarkers** |  |  |
| NYHA class | 282 | 2 (2, 3) |
| Troponin T (ng/mL) | 263 | 47 (34, 65) |
| NT-proBNP (pg/mL) | 301 | 1,745 (878, 3,185) |
| eGFR (mL/min/1.73 m<sup>2</sup>) | 295 | 59 (46, 73) |
| CPET watt achieved (W) | 83 | 108 (73, 147) |
| CPET watt performance (%) | 81 | 77 (59, 98) |
| **NAC Disease staging** |  |  |
| NAC stage | 288 | 1 (1, 2) |
| **Concomitant medications** |  |  |
| Furosemide equivalent dose (mg) | 299 | 2.5 (0.0, 5.0) |
| Other diuretics | 298 | 21 (7.0%) |
| MRA | 299 | 62 (21%) |

*Data are presented as median (Q1–Q3) for continuous variables and n (%) for categorical variables.*
*BMI, body mass index; CPET, cardiopulmonary exercise testing; eGFR, estimated glomerular filtration rate; ICD, implantable cardioverter defibrillator; MRA, mineralocorticoid receptor antagonist; NAC, National Amyloidosis Centre; NT-proBNP, N-terminal pro-brain natriuretic peptide; NYHA, New York Heart Association.*

**Table 2.** Baseline echocardiographic parameters.

| Characteristic | N | Median (IQR) or n (%) |
| --- | --- | --- |
| **LV Structure** |  |  |
| IVSd (mm) | 312 | 14 (12, 16) |
| LV mass indexed (g/m<sup>2</sup>) | 285 | 114 (97, 143) |
| LV EDV indexed (mL/m<sup>2</sup>) | 267 | 43 (36, 52) |
| **LV Systolic Function** |  |  |
| LVEF (%) | 277 | 55 (46, 61) |
| LV GLS (%) | 293 | -14.4 (-17.1, -12.0) |
| LVOT VTI (cm) | 259 | 18.2 (15.2, 21.5) |
| LV CO (L/min) | 277 | 3.1 (2.5, 3.8) |
| **LV Diastolic Function** |  |  |
| E/A ratio | 163 | 1.17 (0.84, 1.74) |
| E/e' | 197 | 14 (11, 18) |
| LA reservoir strain (%) | 268 | 12 (8, 18) |
| LA volume indexed (mL/m<sup>2</sup>) | 263 | 39 (31, 47) |
| **RV Structure and Function** |  |  |
| RV area indexed (cm<sup>2</sup>/m<sup>2</sup>) | 278 | 10 (9, 12) |
| RV FAC (%) | 273 | 32 (24, 39) |
| RV FWLS (%) | 97 | -19 (-23, -14) |
| TAPSE (mm) | 269 | 19 (15, 22) |
| RV S' (cm/s) | 246 | 10.6 (8.6, 13.4) |

*Data are presented as median (Q1–Q3) for continuous variables and n (%) for categorical variables.*
*E/e', early diastolic mitral inflow velocity (E) to early diastolic mitral annular velocity (e') ratio; FWLS, free wall longitudinal strain; GLS, global longitudinal strain; ICD, implantable cardioverter defibrillator; IVSd, interventricular septal wall thickness in diastole; LA, left atrial; LV, left ventricular; LVEF, left ventricular ejection fraction; LVOT VTI, left ventricular outflow tract velocity time integral; RV, right ventricular; S', systolic annular velocity; TAPSE, tricuspid annular plane systolic excursion.*

### Univariable Predictors of Outcomes

In univariable Cox regression (**Table 3**), significant predictors included age, coronary heart disease, atrial fibrillation, NYHA class ≥II, and biomarkers including NT-proBNP, troponin T, and eGFR. Among echocardiographic parameters, left ventricular strain, systolic function (LVEF and LVOT VTI), diastolic function (LA reservoir strain, E/e’ and E/A ratio), and right ventricular parameters (TAPSE, RV S’ and RV FAC) demonstrated significant prognostic value. RV free wall longitudinal strain (RV FWLS) did not reach statistical significance in (HR 1.04 [0.98–1.11], p=0.171). Tafamidis treatment (assessed as time-dependent covariate) showed a trend toward reduced mortality.

**Table 3.** Univariable Cox regression.

| Variable | HR | 95% CI | P |
| --- | --- | --- | --- |
| **Demographics and Clinical Variables** |  |  |  |
| Age per 1 year | 1.08 | 1.05–1.11 | < 0.001 |
| Male sex | 0.94 | 0.5–1.77 | 0.860 |
| BMI per 1 kg/m<sup>2</sup> | 0.96 | 0.92–1.01 | 0.113 |
| Coronary heart disease | 1.76 | 1.19–2.6 | 0.004 |
| Atrial fibrillation | 2.75 | 1.82–4.17 | < 0.001 |
| Carpal tunnel syndrome | 0.68 | 0.42–1.09 | 0.111 |
| Spinal stenosis | 1.50 | 0.9–2.48 | 0.117 |
| Pacemaker or ICD | 1.22 | 0.74–2.04 | 0.436 |
| **Functional Status and Biomarkers** |  |  |  |
| NYHA class ≥II | 4.19 | 1.69–10.37 | 0.002 |
| Ln Troponin T per 1 | 2.06 | 1.6–2.64 | < 0.001 |
| Ln NT-proBNP per 1 | 2.70 | 2.13–3.42 | < 0.001 |
| eGFR per 1 mL/min/1.73m<sup>2</sup> | 0.97 | 0.96–0.98 | < 0.001 |
| CPET watt achieved per 10 W | 0.94 | 0.87–1.02 | 0.114 |
| CPET watt performance per 10% | 1.01 | 0.91–1.13 | 0.804 |
| **Medications** |  |  |  |
| Furosemide equivalent dose per 5 mg | 1.17 | 1.11–1.24 | < 0.001 |
| **LV Structure** |  |  |  |
| IVSd per 1 mm | 1.02 | 0.95–1.09 | 0.590 |
| LV mass indexed per 1 g/m<sup>2</sup> | 1.00 | 1–1 | 0.831 |
| LV EDV indexed per 1 mL/m<sup>2</sup> | 1.00 | 1–1 | 0.916 |
| **LV Systolic Function** |  |  |  |
| LVEF per 1% | 0.94 | 0.93–0.96 | < 0.001 |
| LV-GLS per 1% | 1.17 | 1.1–1.24 | < 0.001 |
| LVOT VTI per 1 cm | 0.90 | 0.85–0.95 | < 0.001 |
| **LV Diastolic Function** |  |  |  |
| E/A ratio per 0.1 | 1.07 | 1.02–1.13 | 0.008 |
| E/e' per 1 | 1.05 | 1–1.11 | 0.038 |
| LA reservoir strain per 1% | 0.92 | 0.89–0.96 | < 0.001 |
| LA volume indexed per 1 mL/m<sup>2</sup> | 1.00 | 1–1 | 0.839 |
| **RV Structure and Function** |  |  |  |
| RV area indexed per 1 cm<sup>2</sup>/m<sup>2</sup> | 1.00 | 0.99–1.02 | 0.908 |
| RV FAC per 1% | 0.97 | 0.95–0.99 | 0.009 |
| RV FWLS per 1% | 1.04 | 0.98–1.11 | 0.171 |
| TAPSE per 1 mm | 0.88 | 0.83–0.92 | < 0.001 |
| RV S' per 1 cm/s | 0.81 | 0.75–0.88 | < 0.001 |
| **Treatment** |  |  |  |
| Tafamidis initiation<sup>a</sup> | 0.77 | 0.55–1.09 | 0.140 |

*BMI, body mass index; CI, confidence interval; CPET, cardiopulmonary exercise testing; E/e', early diastolic mitral inflow velocity (E) to early diastolic mitral annular velocity (e') ratio; eGFR, estimated glomerular filtration rate; FAC, fractional area change; FWLS, free wall longitudinal strain; GLS, global longitudinal strain; HR, hazard ratio; ICD, implantable cardioverter defibrillator; IVSd, interventricular septal wall thickness in diastole; LA, left atrial; LV, left ventricular; LVEF, left ventricular ejection fraction; LVOT VTI, left ventricular outflow tract velocity time integral; MRA, mineralocorticoid receptor antagonist; NAC, National Amyloidosis Centre; NT-proBNP, N-terminal pro-brain natriuretic peptide; NYHA, New York Heart Association; RV, right ventricular; S', systolic annular velocity; TAPSE, tricuspid annular plane systolic excursion.*
<sup>a</sup>assessed as time-dependent covariate

### Correlation analysis

Correlation analysis of echocardiographic parameters revealed moderate to strong correlations between several variables (**Supplemental Figure S1**). Strong correlations were observed between LVEF and LV-GLS (r=−0.79), LVEF and RV FWLS (r=−0.69), and LA reservoir strain and LAVI (r=−0.67). These correlations informed variable selection for multivariable analyses, where collinear parameters were not entered simultaneously.

### Multivariable analysis

In this multivariable analysis (**Table 4**), LV-GLS (HR per 1%: 1.13 [1.03–1.25], p = 0.011) and RV FAC (HR per 1%: 0.96 [0.93–0.99], p = 0.014) remained independent predictors of the composite endpoint, while IVSd (p = 0.192) and E/e’ (p = 0.914) did not reach statistical significance after adjustment. Age was independently associated with outcome (HR per 1 year: 1.06 [1.02–1.10], p = 0.001), whereas sex was not (p = 0.082). The C-index of the model was 0.716 [0.642–0.789].

**Table 4.** Multivariable cox regression.

| Variable | HR | 95% CI | P |
| --- | --- | --- | --- |
| Age, per 1 year | 1.06 | 1.02–1.10 | **0.001** |
| Male sex | 0.40 | 0.14–1.12 | 0.082 |
| IVSd, per mm | 0.93 | 0.84–1.04 | 0.192 |
| LV-GLS, per % | 1.13 | 1.03–1.25 | **0.011** |
| E/e', per 1 | 1.00 | 0.94–1.06 | 0.914 |
| RV FAC, per % | 0.96 | 0.93–0.99 | **0.014** |

*HR, hazard ratio; CI, confidence interval; IVSd, interventricular septal thickness in diastole; LV-GLS, left ventricular global longitudinal strain; RV FAC, right ventricular fractional area change.*

### Kaplan-Meier Analysis of Individual Parameters

Kaplan-Meier analysis demonstrated significant prognostic stratification for multiple echocardiographic parameters (**Figure 1**). LV-GLS (cutoffs: −18% and −14%), LVEF (cutoffs: 40% and 52%), and RV FAC (cutoffs: 30% and 35%) each showed strong discrimination (all overall log-rank p<0.001), while IVSd showed no significant stratification (log-rank p=0.661).

**Figure 1.**
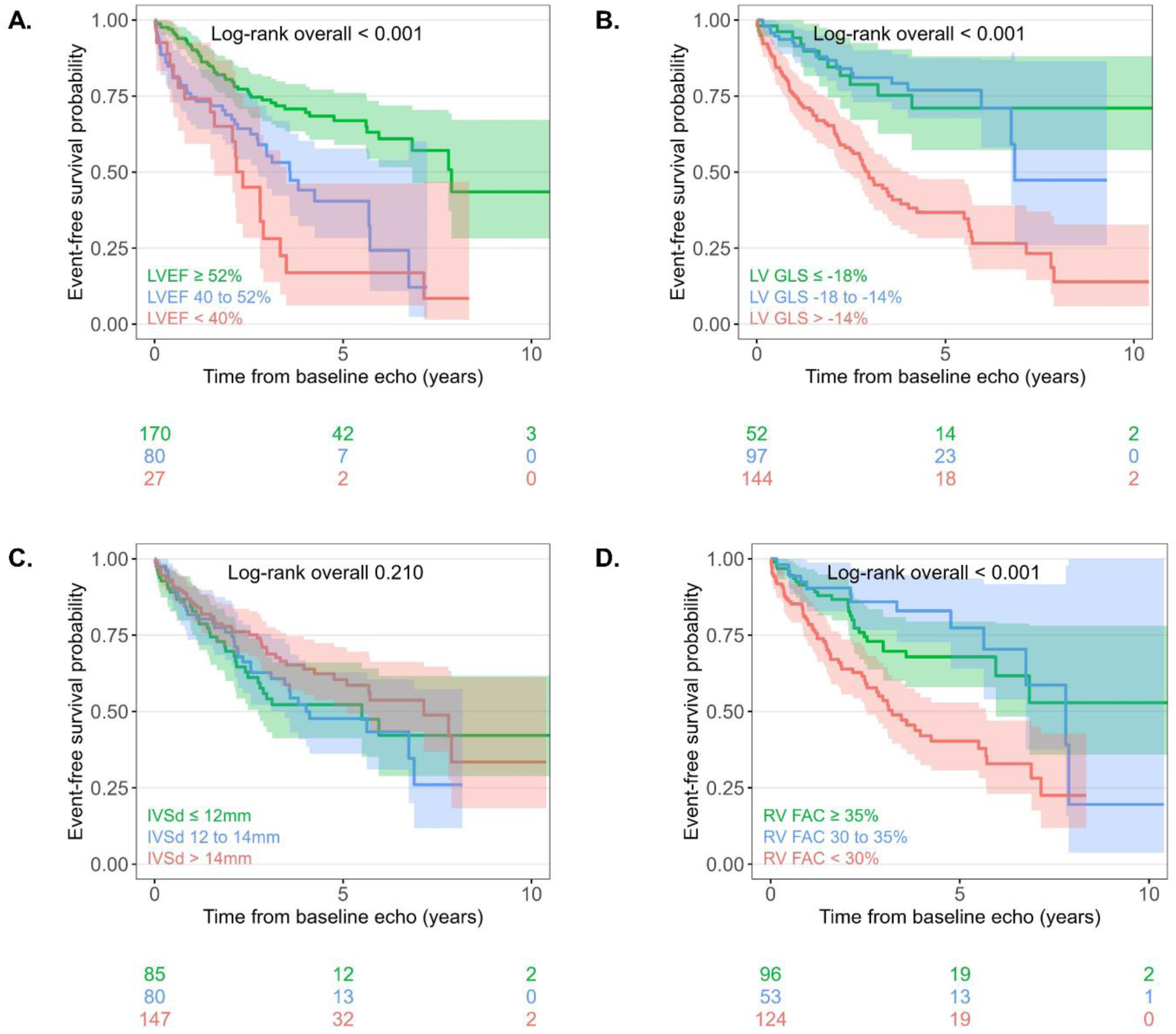
Kaplan-Meier Analysis of Echocardiographic Parameters. Kaplan-Meier curves show event-free survival stratified by AI-derived echocardiographic parameters (A, LVEF; B, LV-GLS; C, IVSd; D, RV FAC). Patients were stratified into three groups per parameter using established clinical cutoffs, with log-rank maximization applied to further stratify abnormal values. IVSd, interventricular septal wall thickness in diastole; LVEF, left ventricular ejection fraction; LV-GLS, left ventricular global longitudinal strain; RV FAC, right ventricular fractional area change.

### Echocardiographic Staging System

A systematic screening of 15 candidate echocardiographic parameters in combination with LV-GLS identified RV FAC as the optimal second parameter (**Supplemental Table S1**). The echo staging combining LV-GLS and RV FAC, stratified patients into low risk (both normal), moderate risk (one abnormal), and high risk (both abnormal) FAC, using cutoffs of −14% for LV-GLS and 30% for RV FAC. This staging system demonstrated strong prognostic stratification (**Figure 2A**) with clear separation of event-free survival curves (log-rank p<0.001). Compared to low-risk patients, moderate-risk patients had a 3-fold increased hazard (95% CI 1.70–5.91, p<0.001), while high-risk patients had a 6-fold increased hazard (95% CI 3.22–10.30, p<0.001). Baseline characteristics differed significantly across echo risk categories (**Table 5**). High-risk patients had a higher prevalence of atrial fibrillation (58% vs. 33% in the low-risk group, p=0.004), markedly elevated NT-proBNP levels (3,039 vs. 862 pg/mL, p<0.001), higher NYHA class (median 2 [IQR 2–3] vs. 2 [IQR 2–2], p<0.001), and higher diuretic requirements (furosemide equivalent dose 2.5 vs. 0.0 mg, p=0.011), confirming that echo staging captures relevant differences in disease severity. The likelihood ratio test indicated echocardiographic staging provided significant incremental prognostic value beyond NAC staging and age (p<0.001; **Figure 2B**), with the C-index improving from 0.730 [95% CI 0.670–0.790] to 0.772 [0.718–0.826] upon addition of echocardiographic staging. Within each NAC stage, echo risk categories identified patients with significantly different annualized event rates (**Figure 3**, NAC Stage I: p=0.002; Stage II: p<0.001; Stage III: p=0.036), demonstrating that echocardiographic staging provides additional prognostic information beyond established disease severity categories.

**Figure 2.**
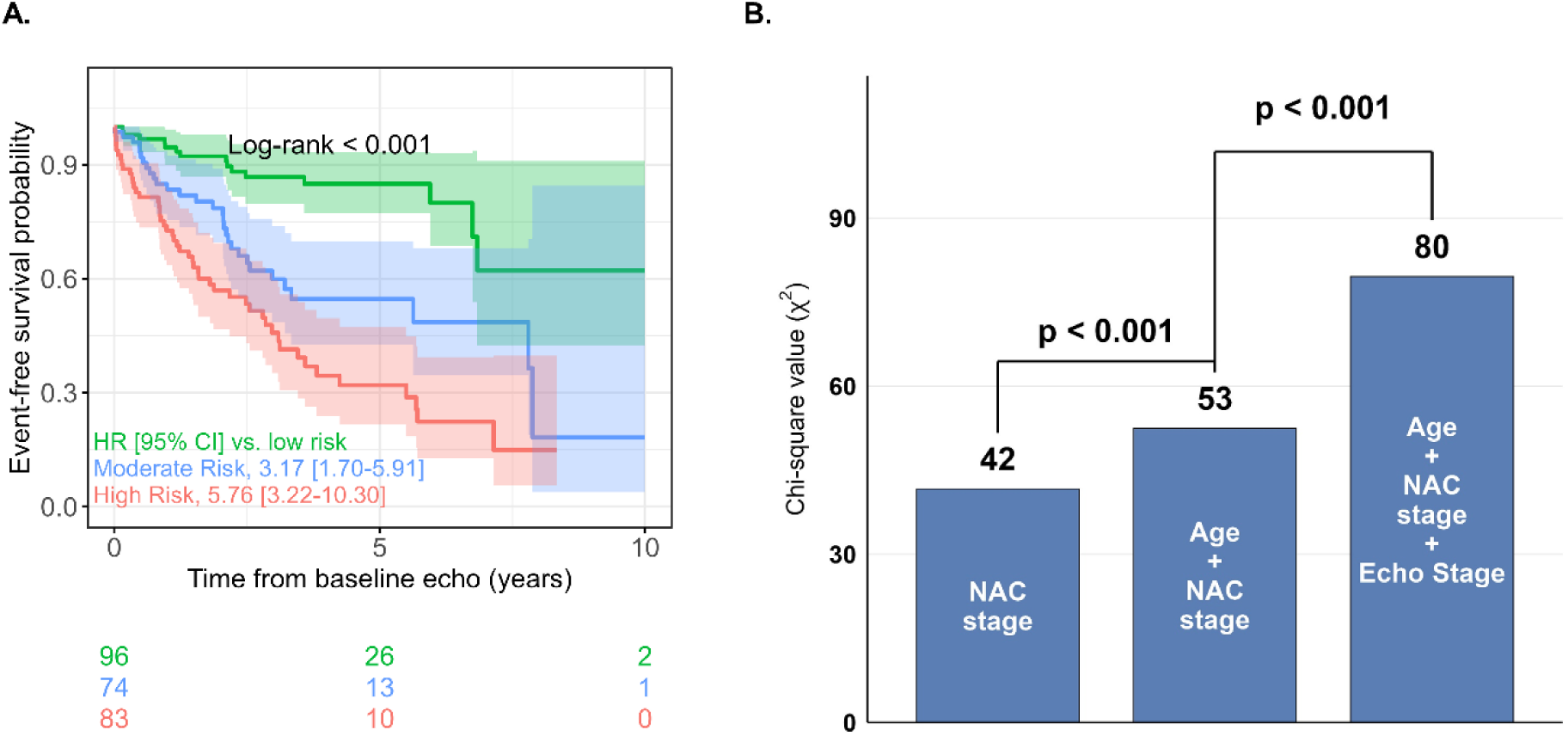
Echo Staging LV-GLS and RV FAC: Event-Free Survival. A, Cumulative event-free survival stratified by echo staging risk groups. B, Likelihood ratio test. LV-GLS, left ventricular global longitudinal strain; NAC, National Amyloidosis Centre; RV FAC, right ventricular fractional area change.

**Figure 3.**
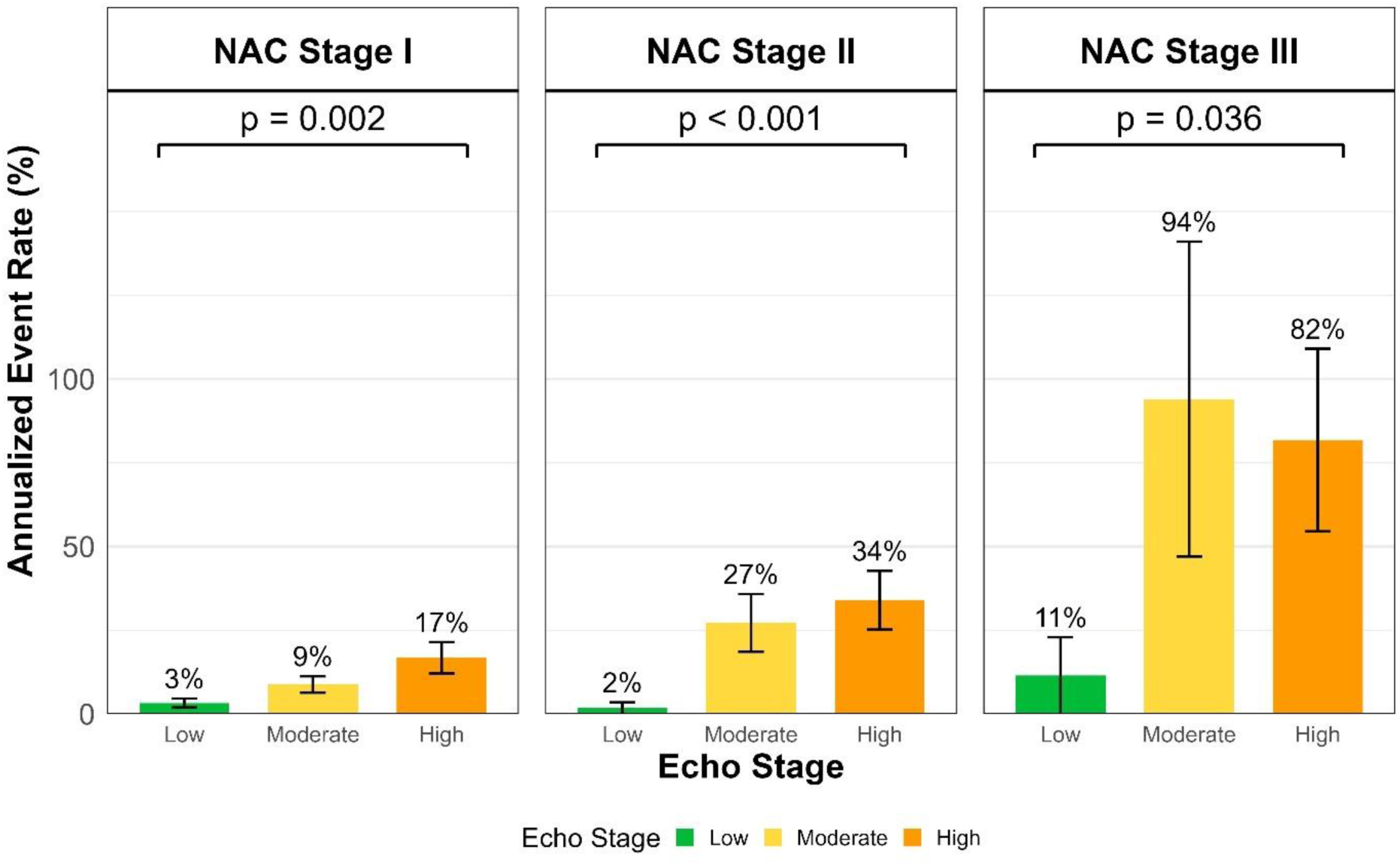
Annualized event rates. The bars represent the annualized event rate for low, moderate, and high-risk groups based on Echo Staging LV-GLS + RV FAC within NAC stages. P-values derived from Poisson regression comparing event rates across echo risk groups within each NAC stage. LV-GLS, left ventricular global longitudinal strain; NAC, National Amyloidosis Centre; RV FAC, right ventricular fractional area change.

**Table 5.**
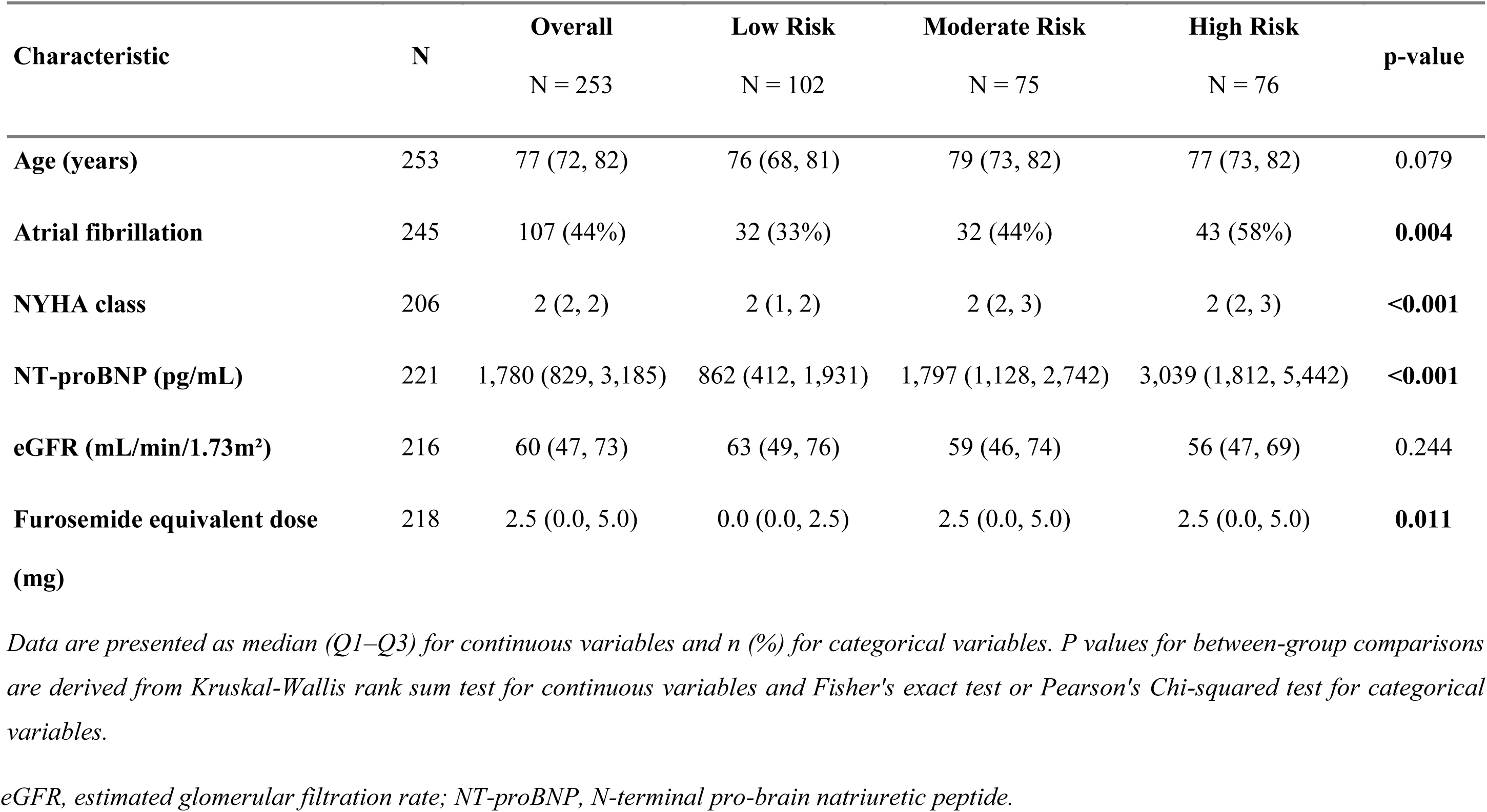
Baseline characteristics by echo staging combining LV-GLS and RV FAC.

| Characteristic | N | Overall<br>N = 253 | Low Risk<br>N = 102 | Moderate Risk<br>N = 75 | High Risk<br>N = 76 | p-value |
| --- | --- | --- | --- | --- | --- | --- |
| Age (years) | 253 | 77 (72, 82) | 76 (68, 81) | 79 (73, 82) | 77 (73, 82) | 0.079 |
| Atrial fibrillation | 245 | 107 (44%) | 32 (33%) | 32 (44%) | 43 (58%) | **0.004** |
| NYHA class | 206 | 2 (2, 2) | 2 (1, 2) | 2 (2, 3) | 2 (2, 3) | **<0.001** |
| NT-proBNP (pg/mL) | 221 | 1,780 (829, 3,185) | 862 (412, 1,931) | 1,797 (1,128, 2,742) | 3,039 (1,812, 5,442) | **<0.001** |
| eGFR (mL/min/1.73m<sup>2</sup>) | 216 | 60 (47, 73) | 63 (49, 76) | 59 (46, 74) | 56 (47, 69) | 0.244 |
| Furosemide equivalent dose<br>(mg) | 218 | 2.5 (0.0, 5.0) | 0.0 (0.0, 2.5) | 2.5 (0.0, 5.0) | 2.5 (0.0, 5.0) | **0.011** |

Data are presented as median (Q1–Q3) for continuous variables and n (%) for categorical variables. P values for between-group comparisons are derived from Kruskal-Wallis rank sum test for continuous variables and Fisher's exact test or Pearson's Chi-squared test for categorical variables.
eGFR, estimated glomerular filtration rate; NT-proBNP, N-terminal pro-brain natriuretic peptide.

### Subgroup Analysis in Tafamidis-Treated Patients

Restricting the analysis to tafamidis-treated patients (n=177), the echo risk staging system retained strong prognostic discrimination (log-rank p < 0.001; C-index 0.752 [0.679–0.825]), with high-risk patients demonstrating a 4.5-fold increased hazard compared to low-risk patients after adjustment for age and NAC stage (HR 4.54 [1.90–10.81], p < 0.001; **Supplemental Figure S2**), confirming that echo-derived risk stratification provides prognostic information independent of tafamidis treatment status.

### Performance AI versus Human Measurements

Time-dependent AUC analysis demonstrated moderate-to-good discriminative performance across AI-derived echocardiographic parameters at 1 year, with time-dependent AUC values ranging from 0.548 to 0.703 (**Supplemental Table S2**). Discriminative performance remained largely stable at 2 years. ROC curves for 1-year event prediction demonstrated comparable performance between AI-derived and human-measured parameters (**Figure 4**). For LVEF, AI-derived measurements achieved an AUC of 0.66 [0.55–0.77] compared to 0.69 [0.59–0.80] for human measurements (ΔAUC = −0.03, p = 0.40). For LV-GLS, AI-derived AUC was 0.64 [0.52–0.77] versus 0.62 [0.52–0.73] for human measurements (ΔAUC = 0.02, p = 0.65). All other parameters tested (E/e’, LAVI, TAPSE, RV S’, **Supplemental Table S3**) similarly showed no statistically significant differences in AUC (all p>0.05), indicating comparable prognostic performance.

**Figure 4.**
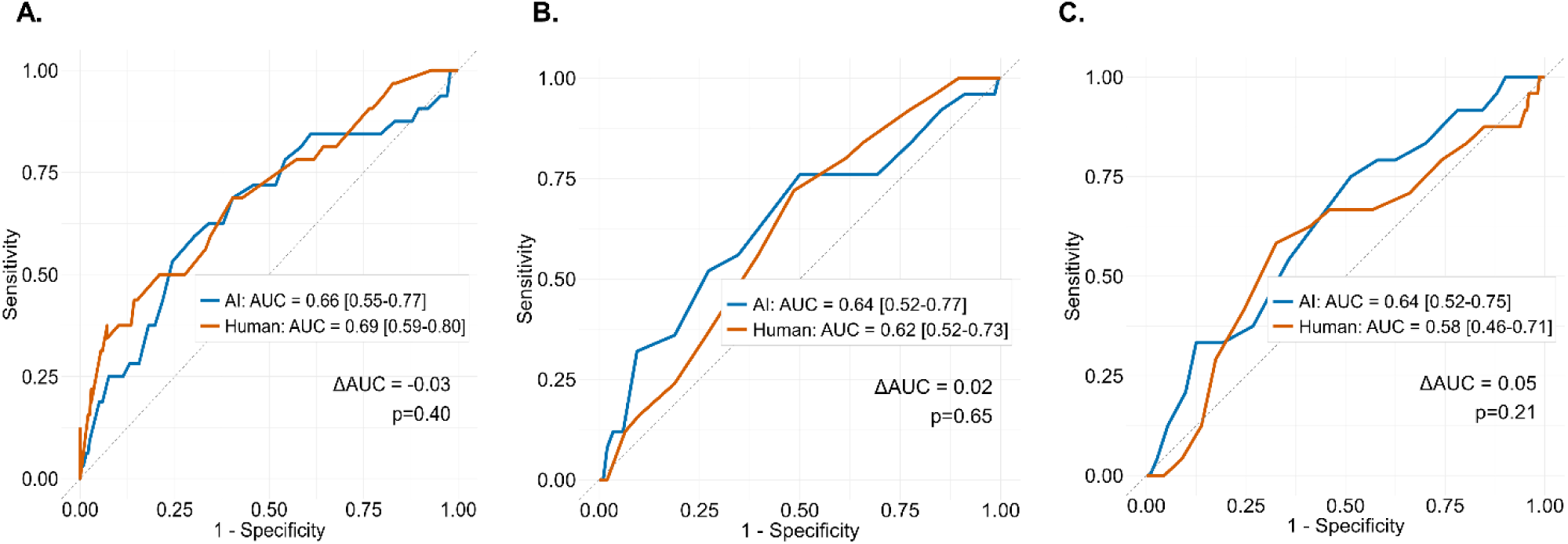
AI-Derived vs. Human-Measured Parameters for 1-Year Event Prediction. Area under the curve (AUC) values with 95% confidence intervals and DeLong p-values for pairwise comparison are shown for LVEF (A), LV-GLS (B) and TAPSE (C). AUC, area under the curve; LV-GLS, left ventricular global longitudinal strain; LVEF, left ventricular ejection fraction; ROC, receiver operating characteristic; TAPSE, tricuspid annular plane systolic excursion.

## DISCUSSION

The present study demonstrates the independent and incremental prognostic value of AI-derived echocardiographic measurements at time of diagnosis beyond established biomarker-based NAC disease staging in cardiac transthyretin amyloidosis. A simple two-parameter staging system combining LV-GLS and RV FAC successfully stratified patients into distinct risk categories, with high-risk patients experiencing a 6-fold increased hazard compared to low-risk patients. Importantly, echocardiographic staging provided independent prognostic information within the NAC stages, with annualized event rates increasing progressively across echo risk categories and revealing considerable heterogeneity within traditional staging system. Furthermore, AI-derived echocardiographic measurements demonstrated comparable prognostic performance to human measurements supporting their clinical utility for risk stratification.

### Comparison to the Literature

Current disease staging in ATTR-CM is primarily based on biomarkers. However, recent evidence suggests that incorporating echocardiographic parameters, particularly LV-GLS, can improve risk stratification.^2^ For ATTRwt-CM, Debonnaire et al. demonstrated that GLS provided prognostic value independent of baseline NAC stage, and remained highly prognostic in tafamidis-treated patients.^2^ Our study confirms and extends these findings to AI-based measurements across all amyloidosis subtypes. In multivariable analysis adjusting for age, sex, and other echocardiographic parameters, both LV-GLS and RV FAC, remained independent prognostic factors.

LV-GLS has been established as a robust prognostic marker in cardiac amyloidosis across multiple studies.^4–6^ Beyond LV-GLS, our systematic screening approach identified RV FAC as the optimal complement to LV-GLS for prognostic staging, highlighting the importance of right ventricular function as an additional prognostic factor in ATTR-CM. This is in line with several prior studies which have demonstrated that right ventricular involvement is a key determinant of prognosis in cardiac ATTR amyloidosis.^7–11^ Several right ventricular parameters have been proposed as prognostic markers in ATTR-CM. Bodez et al. demonstrated the independent prognostic value of TAPSE.^7^ Istratoaie et al. showed that right ventricular free wall strain (RV FWS), assessed by speckle-tracking echocardiography, is an independent prognostic marker for cardiac mortality and heart failure hospitalization, concluding that the ratio of RV FWS to pulmonary arterial systolic pressure (RV FWS/PASp) should be integrated into regular clinical practice for a better evaluation and follow-up of ATTR patients.^8^ In a subsequent analysis, the same group confirmed that among all echocardiographic parameters, RV FWS holds unique prognostic value beyond the current biomarker-based staging system.^9^ In the present study, RV FWLS did not reach statistical significance in univariable analysis, which is likely attributable to the substantially reduced availability of this parameter in our cohort (n=97 of 347 patients), limiting statistical power rather than reflecting an absence of prognostic relevance. Together, prior evidence and the present findings underscore that right ventricular involvement is a key determinant of prognosis in cardiac amyloidosis, likely reflecting the progression of restrictive physiology, elevated filling pressures, and secondary right heart failure. Accordingly, incorporating right-sided parameters into risk stratification algorithms may help to better quantify disease progression.

Our echo staging system incorporating LV-GLS and RV FAC demonstrated a significant increase in annualized event rates across all risk categories, consistent with Debonnaire et al., where GLS provided further prognostic discriminatory power for all-cause mortality within each NAC disease stage stratification. Of note, within NAC stage III, annualized event rates were similarly high in the moderate and high echo risk groups (94% vs. 82%), likely reflecting a ceiling effect in this already high-risk population where the additional discriminatory capacity of echocardiographic staging is inherently limited. Our study revealed a slightly different cutoff for LV-GLS from the one reported by Debonnaire et al. For LV-GLS, we identified a threshold of −14.0%, while Debonnaire et al. suggested a cutoff of −12.8%.^2^ However, our cutoff value for LV-GLS was optimized for a combination of two parameters rather than as isolated parameters, limiting direct comparison between the studies.

Tafamidis showed a non-significant trend toward prognostic benefit in univariable analysis, which is why we did not include it as a covariate in multivariable analyses. However, when analyzing death as a primary outcome, tafamidis showed significance. It is important to note our study was not designed to meet this objective, since the treatment was initiated before baseline for some patients and after baseline for others. Despite this, our echo staging retained strong prognostic discrimination in the subgroup of tafamidis-treated patients, consistent with Debonnaire et al., who demonstrated that GLS retained prognostic value specifically in tafamidis-treated patients.^2^ These findings suggest that echocardiographic risk stratification provides prognostic information independent of treatment effects.

### Comparison AI versus human measurements

AI-derived echocardiographic measurements have been shown to have comparable or superior prognostic performance to human measurements in other diseases. In a large international cohort of patients with acute COVID-19, AI-derived LVEF and GLS were independent predictors of mortality, whereas expert manual measurements were not.^12^ Likewise, in patients with preserved ejection fraction, both AI-derived and manually-derived diastolic function grading demonstrated independent prognostic value, while prior clinical grading lost its significance after adjustment for risk factors.^13^ In the present study, we extend these findings to ATTR-CM, demonstrating that AI-derived measurements achieve comparable prognostic performance to human echocardiographic measurements. Our AUC values for LV GLS for predicting 1-year event-free survival were similar to those reported in previous literature for 1-year mortality prediction.^2^

### Clinical implications

Our findings have several important clinical implications. First, the differences in annualized event rates within biomarker stages underscore substantial clinical heterogeneity within the traditional disease stages. Patients classified in the same NAC stage can have markedly different prognoses. For instance, NAC stage II patients stratified as low versus high echo risk had annualized event rates of 2% versus 34%, respectively. Echocardiography has the potential to capture this gap and further stratify patients. Second, the two-parameter staging system that we suggest, is easily implementable in clinical practice. Most patients with cardiac amyloidosis undergo routine echocardiography at the time of diagnosis and LV-GLS and RV FAC are routinely measured. Third, AI-based measurements have the potential to estimate prognosis not only based on baseline echo, but also based on changes over time. As already shown, a ≥5% decrease of AI-derived LVOT-VTI over 12 months, is independently associated with worse outcome in ATTR-CM.^14^ Finally, in the current era of expanding disease-modifying therapies, accurate baseline risk stratification of the disease is crucial to guide treatment selection: properly staging ATTR patients may help identify patients who are likely to benefit most from effective but costly therapies, enable an assessment of the futility of treatment in patients with advanced disease.

### Limitations

This study has several limitations. First, it is an observational, retrospective study and carries all of the inherent limitations of such a study design. Second, the sensitivity analysis in tafamidis-treated patients is only exploratory in nature since tafamidis treatment was not randomized and sicker patients may have been less likely to receive or initiate therapy. Third, the optimal cutoff values for LV-GLS and RV FAC were derived from the present cohort, and their generalizability to other populations requires prospective validation. Nevertheless, our study also has notable strengths. It extends the assessment of prognostic value to AI-derived measurements with direct comparison to human raters across multiple parameters.

## CONCLUSION

In patients with cardiac amyloidosis, a simple two-parameter echocardiographic staging system combining LV-GLS and RV FAC provides independent and incremental prognostic information beyond established biomarker-based staging. AI-derived echocardiographic measurements demonstrate comparable prognostic performance to human measurements, supporting the feasibility of automated echocardiographic measurements to be sufficiently valid and robust to contribute to risk stratification beyond clinical staging alone. Our findings suggest that routine integration of echocardiographic parameters could refine existing staging systems and inform clinical decision-making in cardiac amyloidosis.

## PERSPECTIVES

### COMPETENCY IN MEDICAL KNOWLEDGE

AI-derived echocardiographic measurements provide independent and incremental prognostic information beyond established biomarker-based staging in ATTR-CM, with performance comparable to human measurements, supporting their integration into routine risk stratification.

### COMPETENCY IN PATIENT CARE AND PROCEDURAL SKILLS

A simple two-parameter staging system combining AI-derived LV-GLS and RV FAC identifies patients with ATTR-CM at up to 6-fold different risk of death or heart failure hospitalization within each biomarker stage, capturing prognostic heterogeneity that established staging alone cannot detect.

### TRANSLATIONAL OUTLOOK

Prospective validation of AI-derived echocardiographic staging in independent cohorts is warranted. In the era of expanding disease-modifying therapies, scalable automated risk stratification holds promise to guide individualized treatment decisions in ATTR-CM.

## Data Availability

The data referred to in that manuscript is not publicly available.

## Abbreviation List

ATTR-CM: transthyretin cardiomyopathy
AUC: area under the curve
E/e’: early diastolic mitral inflow velocity to early diastolic mitral annular velocity ratio
HR: hazard ratio
LV-GLS: left ventricular global longitudinal strain
LVEF: left ventricular ejection fraction
NAC: National Amyloidosis Centre
NT-proBNP: N-terminal pro-brain natriuretic peptide
RV FAC: right ventricular fractional area change
TAPSE: tricuspid annular plane systolic excursion

