## Supplemental Material for "Prognostic value of artificial intelligence-derived echocardiographic measurements in transthyretin cardiomyopathy"

SUPPLEMENTAL APPENDIX

**Figure S1**. Correlation matrix for echocardiographic parameters.

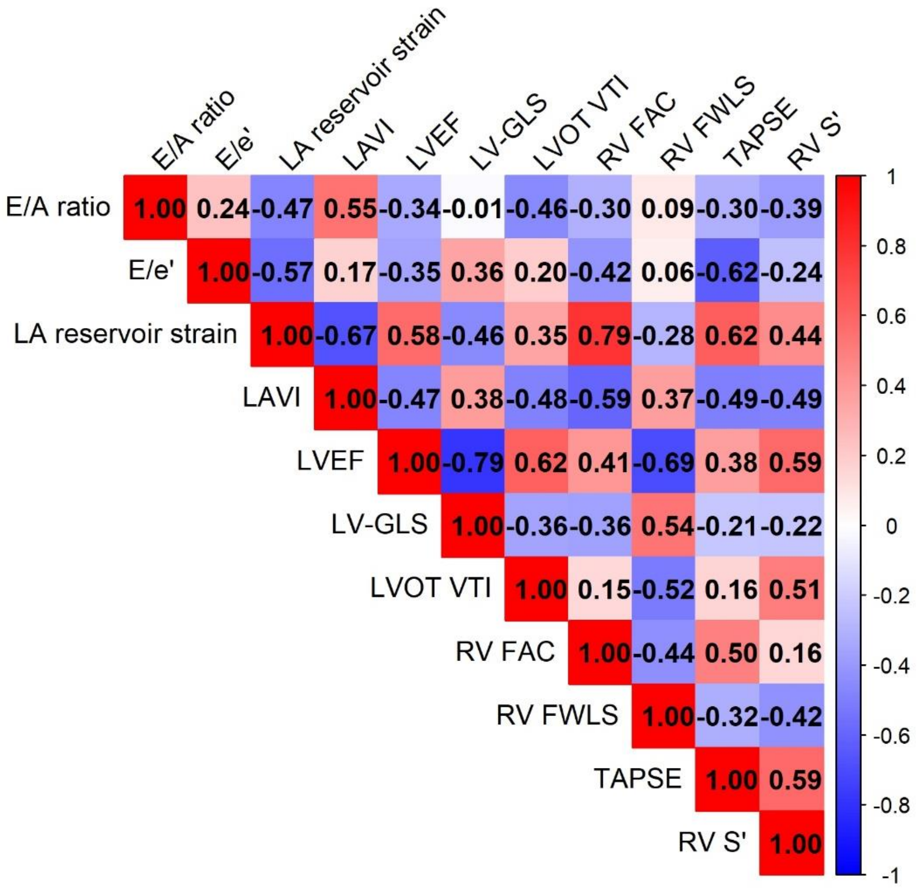

*Pearson correlation coefficients are shown. Color intensity reflects the magnitude of the correlation. E/A, early to late diastolic inflow ratio; E/e', mitral inflow to annular velocity ratio; FAC, fractional area change; FWLS, free wall longitudinal strain; GLS, global longitudinal strain; LA, left atrial; LAVI, left atrial volume index; LV, left ventricular; LVEF, left ventricular ejection fraction; LVOT VTI, left ventricular outflow tract velocity time integral; RV, right ventricular; S', systolic annular velocity; TAPSE, tricuspid annular plane systolic excursion.*

**Figure S2.** Echo Staging in the Tafamidis-Treated Subgroup.

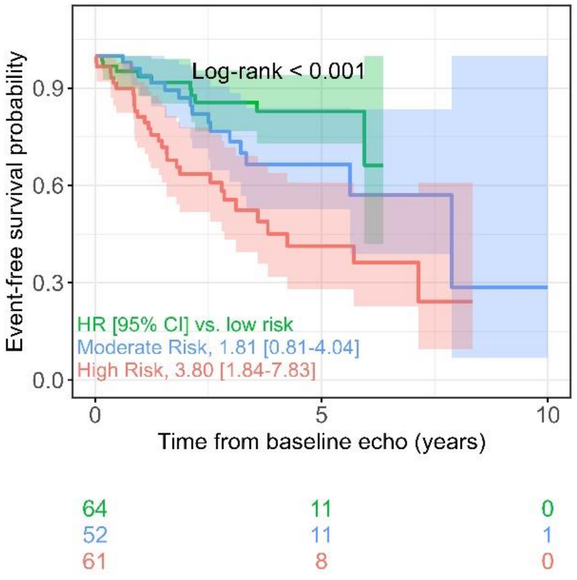

*Cumulative event-free survival stratified by echo staging combining LV-GLS and RV FAC in the tafamidis-treated subgroup.*

*LV-GLS, left ventricular global longitudinal strain; RV FAC, right ventricular fractional area change.*

**Table S1.** Parameter screening for selection of optimal echocardiographic staging system.

| Rank | Parameter | N | Events | $\Delta\chi^2$ | ΔC-index | p (incremental) | Mean $\chi^2$ (stages) | Score |
|---|---|---|---|---|---|---|---|---|
| 1 | RV FAC | 253 | 94 | 27.1 | 0.04 | < 0.001 | 10.44 | 89.72 |
| 2 | LVOT-VTI | 239 | 85 | 24.2 | 0.04 | < 0.001 | 8.40 | 77.10 |
| 3 | TAPSE | 252 | 92 | 21.7 | 0.03 | < 0.001 | 10.08 | 77.04 |
| 4 | RV S' | 233 | 86 | 18.7 | 0.04 | < 0.001 | 10.38 | 72.24 |
| 5 | RV FWLS | 89 | 26 | 11.1 | 0.12 | 0.003 | 12.51 | 71.83 |
| 6 | LVEF | 268 | 106 | 20.6 | 0.02 | < 0.001 | 8.35 | 68.65 |
| 7 | RVEDAI | 258 | 98 | 17.9 | 0.03 | < 0.001 | 9.04 | 66.22 |
| 8 | LA Reservoir | 259 | 101 | 15.9 | 0.02 | < 0.001 | 8.59 | 59.27 |
| 9 | E/A | 156 | 48 | 14.2 | 0.01 | 0.001 | 8.66 | 55.48 |
| 10 | LVEDVI | 259 | 102 | 13.8 | 0.02 | < 0.001 | 7.80 | 52.70 |
| 11 | LAVI | 254 | 96 | 13.7 | 0.02 | < 0.001 | 6.80 | 50.00 |
| 12 | LVCO | 268 | 106 | 11.5 | 0.01 | 0.002 | 7.02 | 45.26 |
| 13 | E/e' | 190 | 64 | 6.1 | 0.02 | 0.029 | 7.68 | 37.64 |
| 14 | LVMI | 262 | 103 | 6.3 | 0.01 | 0.023 | 4.72 | 28.26 |
| 15 | IVSd | 285 | 109 | 1.1 | 0.00 | 0.271 | 2.40 | 9.90 |

**Composite Score = ($\Delta\chi^2 \times 2$) + (Mean $\chi^2$ stages $\times$ 3) + ($\Delta$C-index $\times$ 100)**
$\Delta\chi^2$: Incremental chi-square (Model C vs Model B)
$\Delta$C-index: Incremental C-index improvement
Mean $\chi^2$ (stages): Average log-rank chi-square across NAC stages I-III

*E/A, early to late diastolic inflow ratio; E/e', mitral inflow to annular velocity ratio; FAC, fractional area change; FWLS, free wall longitudinal strain; GLS, global longitudinal strain; LA, left atrial; LAVI, left atrial volume index; LV, left ventricular; LVEF, left ventricular ejection fraction; LVOT VTI, left ventricular outflow tract velocity time integral; RV, right ventricular; S', systolic annular velocity; TAPSE, tricuspid annular plane systolic excursion.*

**Table S2.** Time-dependent discriminative performance of AI-derived echocardiographic parameters

| Parameter | N | Events | 1-year tdAUC | 95% CI | 2-year tdAUC | 95% CI |
|---|---|---|---|---|---|---|
| LVEF | 274 | 109 | 0.676 | 0.592–0.761 | 0.611 | 0.530–0.691 |
| LV-GLS | 290 | 114 | 0.699 | 0.615–0.784 | 0.631 | 0.550–0.711 |
| E/e' | 195 | 65 | 0.548 | 0.420–0.677 | 0.534 | 0.428–0.640 |
| LAVI | 260 | 99 | 0.519 | 0.412–0.625 | 0.540 | 0.450–0.630 |
| RV FAC | 271 | 102 | 0.641 | 0.548–0.734 | 0.688 | 0.607–0.769 |
| TAPSE | 266 | 98 | 0.703 | 0.607–0.799 | 0.690 | 0.600–0.780 |
| RV S' | 244 | 90 | 0.633 | 0.534–0.733 | 0.622 | 0.535–0.709 |

*E/e', mitral inflow to annular velocity ratio; FAC, fractional area change; GLS, global longitudinal strain; LA, left atrial; LAVI, left atrial volume index; LV, left ventricular; LVEF, left ventricular ejection fraction; RV, right ventricular; S', systolic annular velocity; TAPSE, tricuspid annular plane systolic excursion; tdAUC, time-dependent area under the receiver operating characteristic curve.*

**Table S3.** Area under the curve values (AUC) for 1-year event-free survival prediction.

| Parameter | AI_AUC | Human_AUC | Delta_AUC | P |
|---|---|---|---|---|
| LVEF | 0.66 (0.55, 0.77) | 0.69 (0.59, 0.80) | -0.03 | 0.40 |
| LV-GLS | 0.64 (0.52, 0.77) | 0.62 (0.52, 0.73) | 0.02 | 0.65 |
| E/e' | 0.74 (0.59, 0.90) | 0.72 (0.58, 0.87) | 0.02 | 0.81 |
| LAVI | 0.57 (0.47, 0.68) | 0.56 (0.46, 0.67) | 0.01 | 0.84 |
| TAPSE | 0.64 (0.52, 0.75) | 0.58 (0.46, 0.71) | 0.05 | 0.21 |
| RV S' | 0.58 (0.46, 0.71) | 0.63 (0.52, 0.75) | -0.05 | 0.06 |

*E/e', mitral inflow to annular velocity ratio; GLS, global longitudinal strain; LA, left atrial; LAVI, left atrial volume index; LV, left ventricular; LVEF, left ventricular ejection fraction; RV, right ventricular; S', systolic annular velocity; TAPSE, tricuspid annular plane systolic excursion.*
